# Prolonged viral RNA shedding after COVID-19 symptom resolution in older convalescent plasma donors

**DOI:** 10.1101/2020.05.07.20090621

**Authors:** William R Hartman, Aaron S Hess, Joseph Connor

**Affiliations:** Department of Anesthesiology, University of Wisconsin-Madison School of Medicine and Public Health; Department Pathology and Laboratory Medicine, University of Wisconsin-Madison School of Medicine and Public Health

**Keywords:** Coronavirus, COVID-19, RNA detection, symptom resolution

## Abstract

**Importance:** The novel coronavirus, SARS-CoV-2, is responsible for a world-wide pandemic. While the medical community understands the mode of viral transmission, less is known about how long viral shedding occurs once viral symptoms have resolved.

**Objective:** To determine how long the SARS-CoV-2 remains detectable following self-reporting of viral symptom resolution.

**Design:** A cohort of 86 previously SARS-CoV-2 positive patients were re-tested for proof of viral recovery by nasal swab and nucleic acid amplification less than 28 days after self-reported symptom resolution. This study was approved by the University of Wisconsin Institutional Review Board.

**Setting:** A tertiary care center in a mid-size city utilizing a drive-through SAR-CoV-2 testing center.

**Participants:** 86 previously confirmed SARS-CoV-2 positive individuals less than 28 days after self-reported resolution of symptoms evaluated as potential donors for COVID-19 convalescent plasma.

**Intervention:** Participants underwent nasopharyngeal sampling and subsequent nucleic acid amplification for SARS-CoV-2 genes.

**Main Outcome:** SARS-CoV-2 RNA in nasopharyngeal secretions detected by rtPCR.

**Results:** 11/86 (13%) previously confirmed SARS-CoV-2 subjects were still positive at a median of 19 days (range 12-24 days) after symptom resolution. Older patients were more likely to be test-positive, and older positive patients had lower rtPCR C_T_ values. Test-positive patients were not significantly different from test-negative patients with respect to days since symptom recovery.

**Conclusions and Resolution:** These results underscore the necessity of testing COVID-19 convalescent plasma donors less than 28 days after symptom resolution and suggests that COVID-19 positive patients may need to remain in quarantine beyond the recommended two weeks following “recovery.”

**Trial Registration:** n/a

## Introduction

SARS-CoV-2 has infected over 3.1 million people worldwide and caused over 210,000 deaths, with over 1 million infections and 58,000 deaths in the United States.^1,2^ Proving resolution of disease is critical for population health, but this is limited by availability and uncertain interpretation of SARS-CoV-2 serology and the uncertain natural history of viral shedding detected by nucleic acid testing (NAT). A study in China found viral RNA in the stool of 17% of symptom-free patients 9-16 days after onset of disease.^3^ Systematic re-testing has been limited in the United States.

## Methods

This study was approved by the University of Wisconsin Institutional Review Board. Previously laboratory-confirmed COVID-19 patients were recruited in collaboration with state public health officials and screened as possible convalescent plasma donors. Eligible participants less than 28 days symptom-free were invited to undergo repeat testing. Nasopharyngeal swabs were collected in viral media by trained nurses. RNA was extracted using the Maxwell RSC kit (Promega, Madison, WI). SARS-CoV-2 N and RdRP genes were amplified using the Allplex™ 2019-nCoV Assay Kit (Seegene, Seoul, ROK) on the QuantStudio 5 rtPCR system (Applied Biosystems, Foster City, CA). *C_T_* values < 40 for N- and/or RdRP were considered positive. Mean *C_T_* for all positive gene was reported. Comparisons between groups were made with parametric and non-parametric statistics, as appropriate, and linear regression used to compare continuous variables. Analyses were performed with commercial statistical software (Enterprise Guide 7.1, SAS, Cary, NC).

## Results

152 potential participants were screened, of which 5 declined, 54 were ineligible, and 93 were included. 86 of 93 completed testing. 11/86 (13%; 95% confidence interval [CI] 7%, 22%) were positive and 75/86 (87%; 95% CI = 80%, 94%) were negative. Median positive CT was 37.3 (interquartile range [IQR] 35.3, 38.6). The range of days since symptom resolution was 12-24 among participants who tested positive and 11-30 among participants who tested negative. Positive and negative participants were not significantly different with respect to days since resolution of COVID-19 symptoms (median 19 days; IQR 14, 22 vs. 18 days; IQR 15, 21; p = 0.771). Positive participants were significantly older than negative participants (mean 54 years; 95% CI 44, 63 vs. 42 years; 95% CI 38, 46; p = 0.024). *C_T_* values were significantly, inversely associated with age (β = −0.04, r^2^ = 0.389, p=0.04). 8/11 positive participants retested negative at a median of 22 days (IQR 19, 28) after resolution of symptoms.

## Discussion

We do not know if or for how long a patient remains contagious after COVID-19 symptoms resolve. Pre-symptomatic patients shed infectious virions.^4^ Previous studies have detected SARS-CoV-2 RNA in respiratory secretions 13-29 days after symptom onset.^5^ Patient isolation measures are dependent on assumptions about duration of viral shedding. We found that among 86 persons with prior laboratory-confirmed COVID-19 screened for a convalescent plasma donation, 13% still had detectable viral RNA in their nasopharyngeal secretions at a mean of 18 days after symptom resolution. Median *C_T_* values indicate that the viral RNA levels are near many commercial assays’ limit of detection.^6^ Positive patients were significantly older than negative patients, and older patients had significantly higher viral RNA loads at time of testing. These data have limitations. Detectable RNA in secretions is not the same as shedding infectious virions, although NAT is the only rapid and widely available method for detecting COVID-19. There are likely biases and confounding, although the nearly identical time-to-test between groups suggests that positive results are not biased by earlier testing. These data demonstrate that COVID-19 patients continue to shed detectable viral RNA for 2-4 weeks after resolution of symptoms. Efficient testing to confirm resolution of COVID-19 for surveillance and convalescent plasma programs should likely focus on patients outside this window.

## Data Availability

Data from this study are not currently available for distribution.

---

**Table 1:** Screening results for previously COVID-19 positive patients recruited to a convalescent plasma program (n=86).

|  | **SARS-CoV-2 Viral RNA PCR Result** |  | **p** |
| --- | --- | --- | --- |
|  | Positive<br>(N = 11) | Negative<br>(N = 75) |  |
| Days since last symptoms<br>*median (interquartile range)* | 19 (14, 22) | 18 (15, 21) | 0.771* |
| Age<br>*mean ± SD* | 53 ± 15 | 43 ± 16 | 0.024† |

\*Wilcoxon-Mann-Whitney test
†T-test

## Notes

### Competing Interest Statement

The authors have declared no competing interest.

### Funding Statement

No external funding was received.

